# Factors associated with delayed umbilical cord clamping in public health facilities in Debremarkos town, northwest Ethiopia

**DOI:** 10.1101/2023.09.20.23295821

**Authors:** Betel Bogale Workineh, Endeshaw Admasu Cherkose, Belayneh Ayanaw Kassie

**Author notes:** **Email adresses:** Betel Bogale Workineh (BBW) Endeshaw Admasu Cherkose (EAC) Belayneh Ayanaw Kassie (BAK). **Corresponding Author:** Betel Bogale Workineh.

## Abstract

**Background:** Delayed cord clamping is a proven intervention to improve both maternal and neonatal health and nutrition. World Health Organization recommends not clamping the umbilical cord before 1 minute of delivery. However, little is known about the timing of umbilical cord clamping, and associated factors in Ethiopia.

**Objective:** To assess the timing of umbilical cord clamping and associated factors among women who gave birth at public health institutions in Debremarkos town, 2022/23.

**Methods:** A facility-based cross-sectional study was conducted from December 01, 2022, to January 30, 2023, among women selected using systematic random sampling. Data was collected through observation and review of medical records using a checklist. The data was entered to Epi-data version 4.6.0.4 and analyzed by STATA 14. Descriptive statistics, bivariable and multivariable logistic regression models were fitted.

**Result:** A study of 388(91.73% response rate) women-newborn pairs found that 206(53.09%) newborns received delayed umbilical cord clamping, with mean and median clamping times of 67.87 ± 39.86 SD and 60s, respectively. In the multivariable analysis, giving birth at the hospital (AOR = 2.47, 95% CI: 1.21-5.03), attended by medical interns (AOR = 2.47, 95% CI: 1.29-5.41), receiving uterotonic for AMTSL at or after 60 seconds of giving birth (AOR = 10.36, 95% CI: 6.02-17.84), Rh-negative mothers (AOR = 3.91, 95% CI: 1.40-10.95), and multiparity (AOR = 0.54, 95% CI: 0.32-0.93) were significantly associated with delayed umbilical cord clamp.

**Conclusion:** In this study, half of the newborns had delayed umbilical cord clamping. However, the result is still unsatisfactory, as the recommendations for delayed cord clamping extend to all newborns who do not require intensive care. Therefore, considering the proven benefit of delayed umbilical cord clamping, obstetric care providers should adhere to clinical guidelines for this proven intervention.

## Introduction

Globally, there were over 1·2 billion cases of iron deficiency anemia(IDA) in 2016 (1). It affects 5% to 25% of preschool children in high-income countries and up to 100% in low-income countries (2). In Ethiopia, its prevalence under-five children is 57%, of which 72% are among infants and young children (3). Also, it has been shown that iron deficiency in infancy contributes to impaired cognitive, motor, and behavioural development that may be irreversible (4,5).

For the first 6 months of life, babies predominantly meet their iron needs from their birth iron endowment, which is influenced by birth weight, gestational age, and the timing of cord clamping (1). There are different policies on the timing of cord clamping; early or immediate cord clamping (ECC) and delayed cord clamping (DCC) (6). The World Health Organization (WHO) defines delayed cord clamping as the clamping of the cord within 1 to 3 minutes of birth (7).

Early cord clamping has been shown to be associated with a double chance of anemia at 3 to 6 months in term infants (8). Likewise, it has been demonstrated that DCC by 60 seconds allows approximately 80 mL of blood to be transferred from the placenta to the newborn and reaches approximately 100 mL by 180 seconds (2). This placental transfusion provides sufficient iron reserves for the first 6–8 months of life, preventing or delaying the development of iron deficiency until other interventions such as the use of iron-fortified foods– can be implemented (7). So, DCC is an evidence-based, low-cost intervention used to prevent IDA and its consequences for children in low- and middle-income countries where its prevalence is high (5).

DCC is also established as a strategy for decreasing rates of intraventricular hemorrhage, necrotizing enterocolitis, and the requirement of transfusion in term and preterm infants (9). A 50% reduction in intraventricular hemorrhage in all grades has been shown in the review of 10 trials (10). In addition, the Cochrane review reported that delayed cord camping may reduce the risk of death before discharge for babies born preterm(11). DCC has been demonstrated to improve oxygen saturation, facilitate spontaneous breathing, and smoother cardiovascular transition (12,13).

As a result of that evidence multiple professional organizations, including the World Health Organization (WHO),the American College of Obstetrics and Gynecology (ACOG) and the National Institutes for Health and Care Excellence, now recommend delayed cord clamping for infants of all gestational ages (4,7,14).

Despite various recommendations, it is observed that there are differences in health care providers’ birth practices and cord clamping times all over the world (15,16). However, in our country, Ethiopia, there is little information regarding the current timing of cord clamping, and there is no study evaluating the factors affecting the timing of cord clamping. Therefore, this study was aimed at assessing the proportion of delayed cord clamping and its associated factors among women who gave birth at public health institutions.

## Methods

### Study design, period, area and population

The institution-based cross-sectional study design was conducted among women who gave birth at public health institutions in Debremarkos town, northwest Ethiopia, from December 01, 2022, to January 30, 2023. Debre Markos town is the administrative town of East Gojjam Zone, which is located 276 km away from Bahir Dar, the capital city of Amhara Regional State, and 300 km away from Addis Ababa (capital city of Ethiopia). Currently, it has seven kebeles (the smallest administrative unit in Ethiopia). Debre Markos town has one comprehensive specialized hospital and four public health centers: Debre Markos, Gozamen, Hidasse, and Wuseta health centers, which provide antenatal, labor, and delivery services.

All laboring women in the public health facility in Debremarkos town were the source of the population. All laboring women in public health facilities in Debremarkos town during the study period were the study population. All laboring women who had had alive fetus and gave birth to an alive newborn were included. Women identified as planning to give birth by caesarean section, mothers whose fetus died during intrapartum, second-born twins, mothers whose new-borns had tight nuchal cords (i.e., wrapped tightly around their necks) and cord clamping done prior to delivery, major fetal anomalies or no intent to resuscitate, and those who arrived at the institution in the expulsive period were excluded.

### Sample size & sampling procedures

The sample size was determined by using the single population proportion formula through StatCalc Epi Info™ version 7.2.4.0, considering the following assumptions: 95% level of confidence, 5% margin of error, and 50% proportion of delayed cord clamping (p-value) since there is no prior study from Ethiopia. The sample size for the first objective was calculated to be 384, and this gave the largest sample size than those calculated for the associated factors. By considering a 10% nonresponse rate, the final sample size was 423.

Systematic random sampling was used to select the study participants from the five public health facilities in Debremarkos town, namely: Debremarkos Comprehensive and Specialized Hospital, Debremarkos Health Center, Gozamen Health Center, Wuseta Health Center, and Hidassie Health Center. The previous three months’ client flow to the five health facilities for delivery service was reviewed from the registration book to estimate the expected number of pregnant women who were admitted for delivery in two months. Therefore, the average number of laboring women admitted to the labor ward in the previous three months was 1061, 60, 54, 46, and 57 for Debremarkos Comprehensive and Specialized Hospital, Debremarkos Health Center, Gozamen Health Center, Wuseta Health Center, and Hidassie Health Center, respectively.

The sample size was proportionally allocated to health centers and the hospital based on the respective health facilities’ three-month deliveries before this study. Accordingly, the proportional allocation to population size for each health facility was 351, 20, 18, 15, and 19 for Debremarkos Comprehensive and Specialized Hospital, Debremarkos Health Center, Gozamen Health Center, Wuseta Health Center, and Hidassie Health Center, respectively. The sampling interval (kth unit) was obtained by dividing the total number of pregnant women who were admitted for delivery in the last two months (calculated to be 852) by the desired sample size (total number of sample sizes). The first woman was randomly chosen for the survey by the lottery method, and then data were collected from all women who gave birth throughout the day and night during the two-month study period. Every kth value (2) according to their sequence of birth was included in the study.

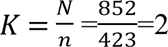

### Operational definitions

#### Delayed cord clamping (DCC)

DCC is referred to as when the interval between the birth of the baby and the time that the first clamp was applied to the umbilical cord is greater than or equal to one minute (7).

### Data collection procedures (instrument, personnel)

For this study, a checklist that was adopted from studies done in different literature was used(12,16,25–29,17–24). Data were collected through participant observation to prevent bias. To complete observations, the data collector used printed checklists to record data and a stopwatch. A stopwatch was used to time the cord clamping interval. The stopwatch was pressed once at the time of the birth and once when the first clamp was applied to the umbilical cord. To prevent bias, we did not define ECC and DCC in the questionnaire but categorized the time points afterward based on the WHO criteria.

Data related to the timing of cord clamping and AMTSL was collected by observation; labor-delivery characteristics, obstetric complications, maternal characteristics, and newborn characteristics were collected by medical record review or observation. There were four midwife data collectors in the hospital (2 for day and 2 for night), eight student midwife data collectors at the health center (one for day and one for night in each health center), and two MSc. midwife supervisors were recruited.

### Data quality control

The training was given on how to collect the data, the way to approach it, observation techniques, how to keep the information, and how to better understand the overall process of the data collection procedure before the actual data collection. And before the actual data collection, a pretest was done on 5% of the sample size at Yejube Primary Hospital to check the language clarity and appropriateness of the checklist. During data collection, the checklist was checked for completeness daily by the supervisor and principal investigator.

### Data processing and analysis

Data were checked, coded, and entered into EPI data version 4.2.0.6; then they were exported to STATA 14 for analysis. The descriptive analysis was done for all variables. Variables were categorized as to whether early cord clamping (ECC) or DCC was used (DCC was defined as ≥ 60 s). Finally, binary logistic regression was used to determine factors of receiving DCC.

Bivariable logistic regression was used to identify statistically significant independent variables, and independent variables having a p-value of less than 0.2 were considered candidates for multivariable logistic regression for controlling confounders (29). Model assumption fulfillment and the multicollinearity test were done prior to multivariate logistic regression. The final logistic model’s goodness of fit was assessed using the Hosmer-Lemeshow goodness of fit test. In multivariable logistic regression, a p-value of <0.05 with a 95% confidence interval for the odds ratio was used to determine the significance of the association.

### Ethical Consideration

Ethical clearance was obtained from the School of Midwifery under the delegation of the University of Gondar institutional review board with ref No. MIDW/30/2015. Official permission was secured from Debremarkos Comprehensive and Specialized Hospital and each health center and submitted to each public health facility department unit focal person.

Healthcare providers were informed that they were going to be observed while attending a birth. Also, they were told that the data would be used only for the study purpose and would not be personally identifiable. In order to maintain the scientific validity of the research, they were not informed that they were specifically observed for their timing of cord clamping. This is supported by the National Research Ethics Review Guideline and the International Ethical Guidelines for Health-related Research Involving Humans. This is due to the fact that this study fulfilled the three conditions set by them to obtain approval for modification or waiver of informed consent. i.e., the data were not personally identifiable; the research has important social value and poses no more than minimal risk to participants (30,31).

Written informed consent, which was prepared in the local language (Amharic), was obtained from the study participants’ mothers. The woman consented at the time of admission to the hospital for labor care when the data collector considered it would be appropriate for her to consent at that time (i.e. if she was not unduly distressed and not in a circumstance where the discussion may have impacted on her ability to cope with labor), and participation in the study was offered at this time. If any concern existed about a woman’s ability to consent, she was excluded from the study.

## Results

### Study population

Among the 421 women who were eligible and consented to have information collected from their births, 388 were included for a measurement of the timing of umbilical cord clamping (**Fig. 1**). Overall, 377 women delivered singletons, and 11 women delivered twins. Second-born twins (n = 11) were excluded. Thus, all subsequent outcomes are reported for the remaining 388 singletons and firstborn twins.

**Fig 1.**
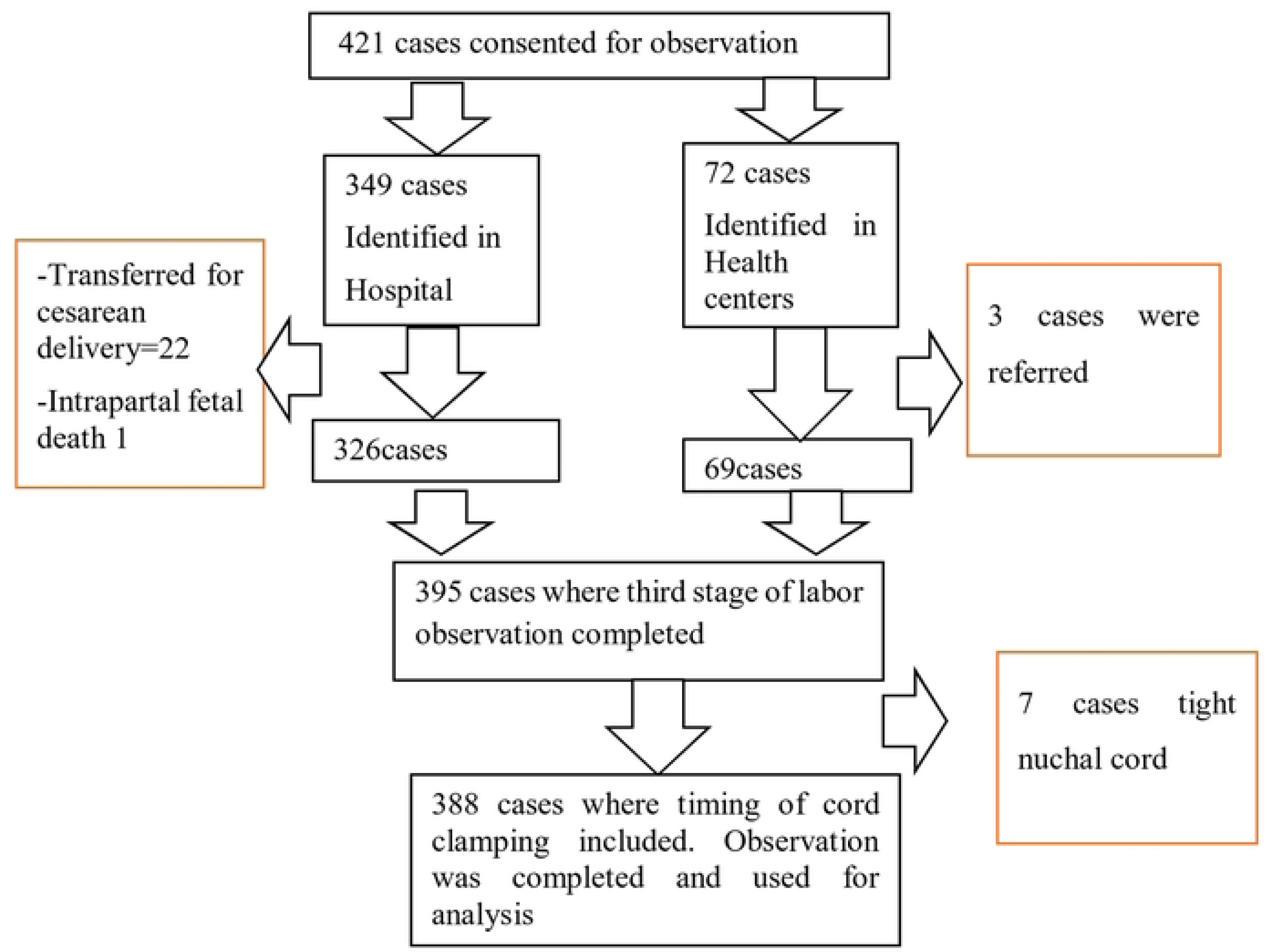
Flow chart of study participants included in this study.

### Maternal characteristics

The mean maternal age in the year was 27.6 (SD of 5.2). The majority (59.79%) of the women were multiparous. Rh-negative mothers were 29 (7.47%). Maternal anemia during pregnancy was found in 5 cases (1.29%), and maternal infection (mothers who were positive for syphilis, HIV, HBV, or HSV) was found in 16 cases (4.12%) (**Table 1**).

**Table 1.**
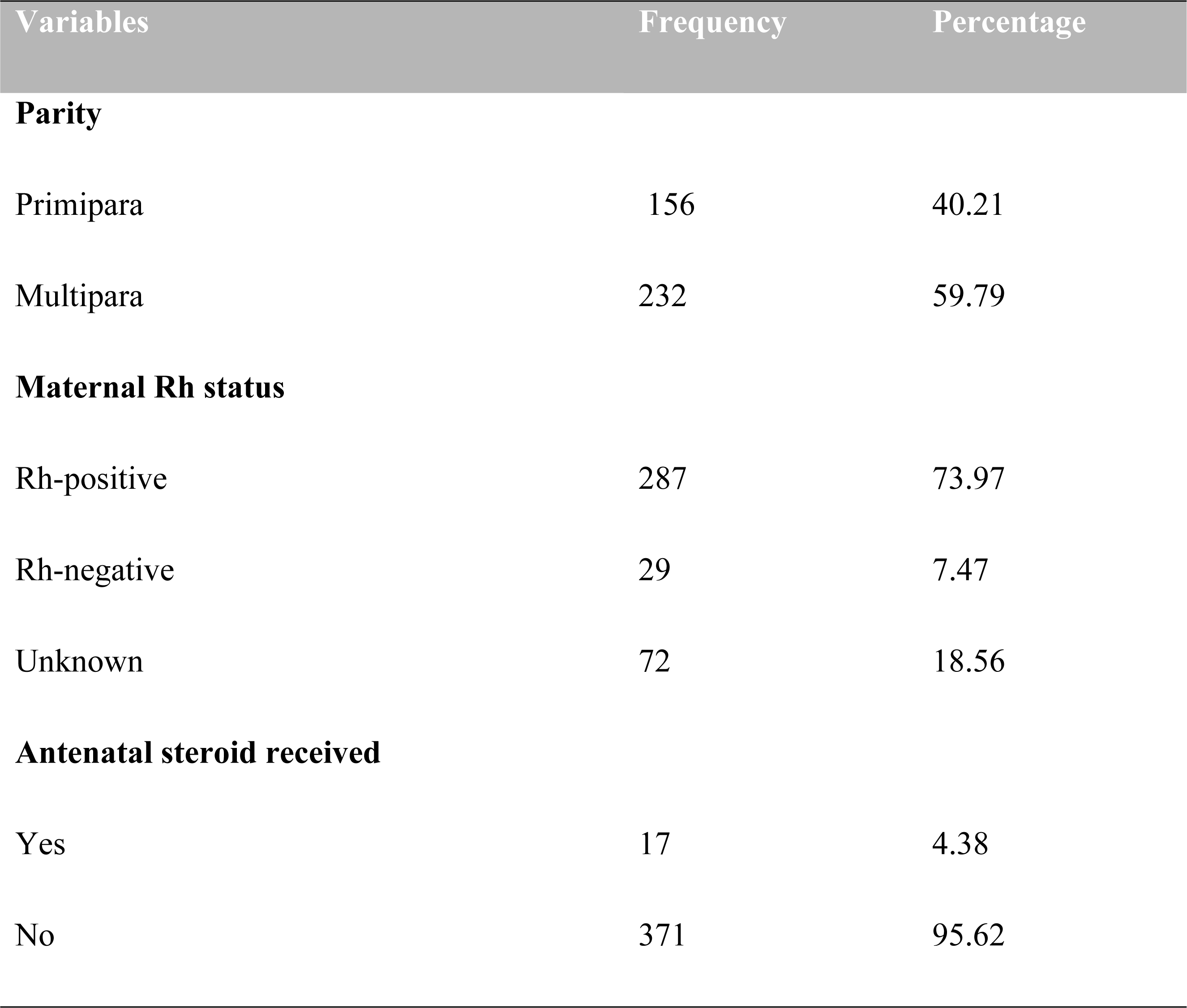
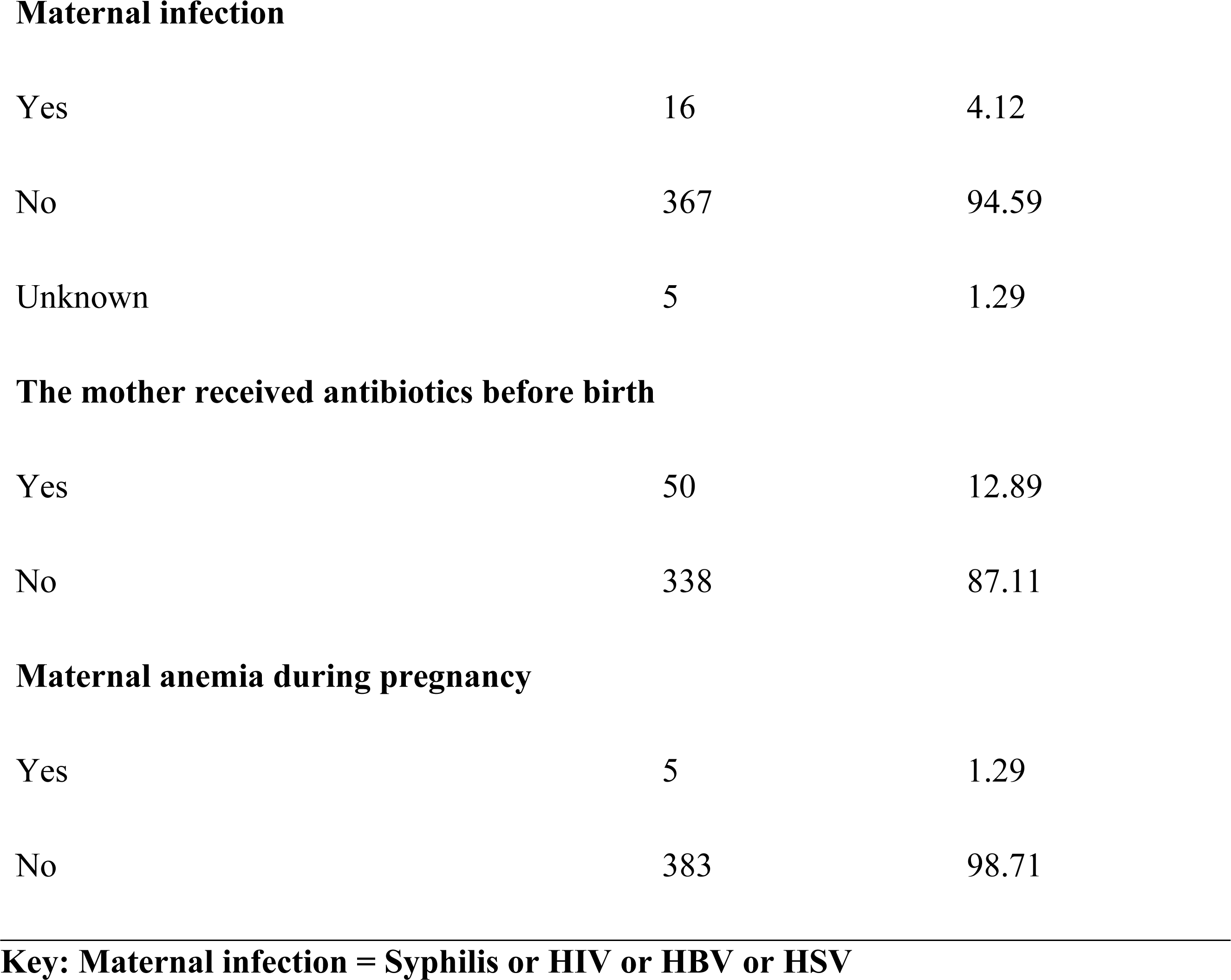
Maternal characteristics among women who gave birth in Debremarkos town public health institutions, north Ethiopia, 2022/23, (n=388)

### Newborn characteristics

Newborns of both sexes had an equal proportion, and the average birth weight was 2941.2g (SD of 446.5). Most of them were in cephalic presentation, while 14 (3.62%) were in breech (**Table 2**). Majority (61.76%) of newborns were born at term. (**Fig. 2**)

Newborn characteristics among women who gave birth in Debremarkos town public health institutions, north Ethiopia, 2022/23, (n=388)

| Variables | Frequency | Percentage |
| --- | --- | --- |
| **Sex of the baby** |  |  |
| Male | 194 | 50.0 |
| Female | 194 | 50.0 |
| **Birth weight** |  |  |
| VLBW (<1500g) | 4 | 1.03 |
| LBW (1500-2500g) | 68 | 17.53 |
| Normal BW (≥2500g) | 316 | 81.44 |
| **Gestation** |  |  |
| Singleton | 377 | 97.16 |
| Multiple (first-born twins) | 11 | 2.84 |
| **Presentation at birth** |  |  |
| Cephalic | 369 | 95.10 |
| Breech | 14 | 3.62 |
| Face | 5 | 2.29 |
| **Apgar at the first minute score <7** |  |  |
| Yes | 83 | 21.39 |
| No | 305 | 78.61 |
| **The newborn needs resuscitation** |  |  |
| Yes | 94 | 24.23 |
| No | 294 | 75.77 |

**Fig 2.**
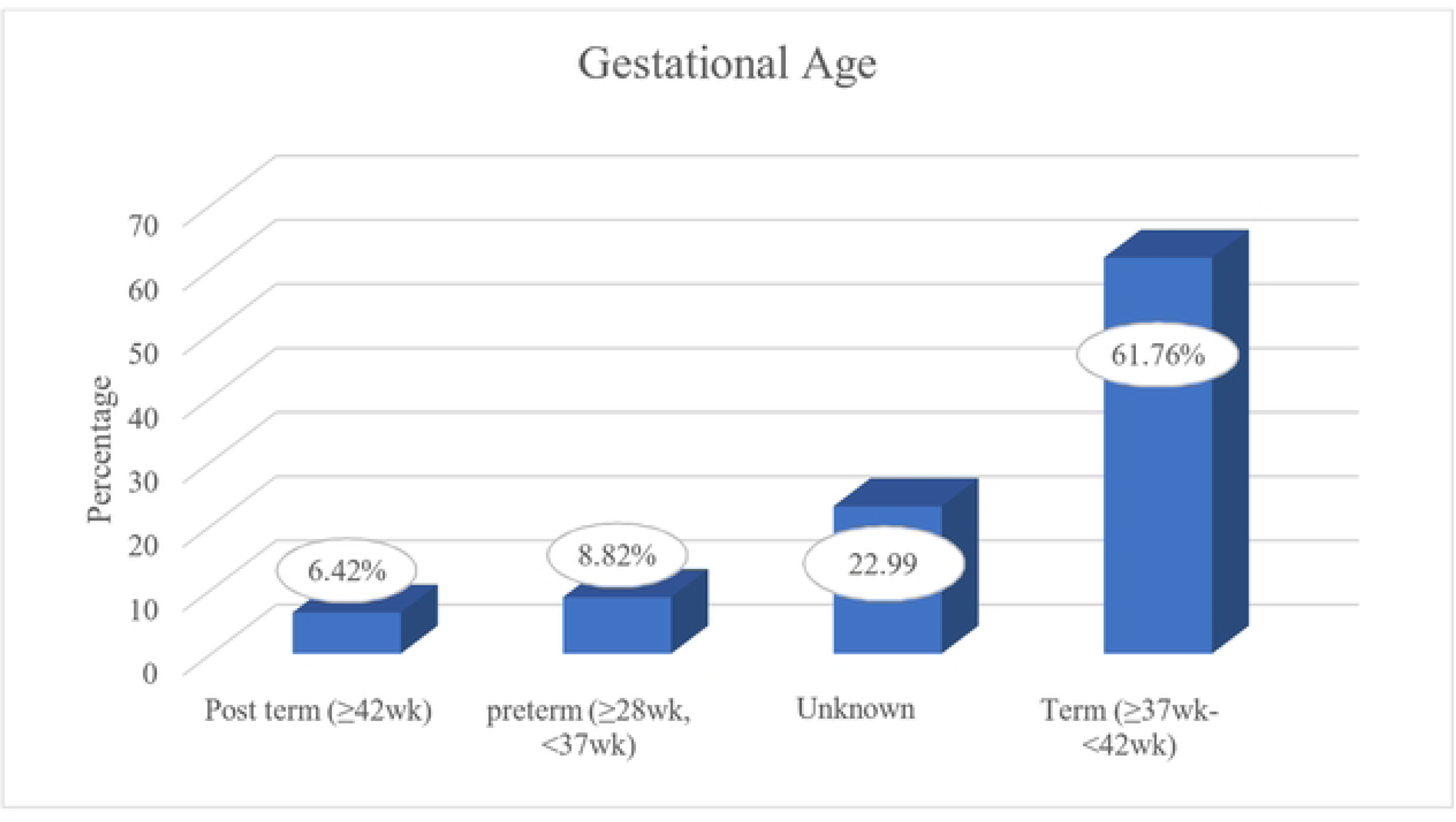
Gestational age a1nong women who gave birth in Debren1arkos town public health institutions, north Ethiopia, 2022/23.

### Labor delivery characteristics

The majority (82.22%) of women gave birth at the hospital, while the rest gave birth in the four health centers. Spontaneous vaginal deliveries were 324 (83.51%), while 54 (13.92%) were instrumental deliveries. More than half (60.82%) of deliveries occur during the day. Metallic clamp forceps were used as an instrument to clamp the umbilical cord in 235 (60.57%) newborns. And cord milking was done in 130 (33.51%) of births (**Table 3**). Intramuscular oxytocin was used as a prophylactic uterotonic during the third stage of labor in all women. The range of uterotonic administration times was from 9 to 351 seconds.

**Table 3.**
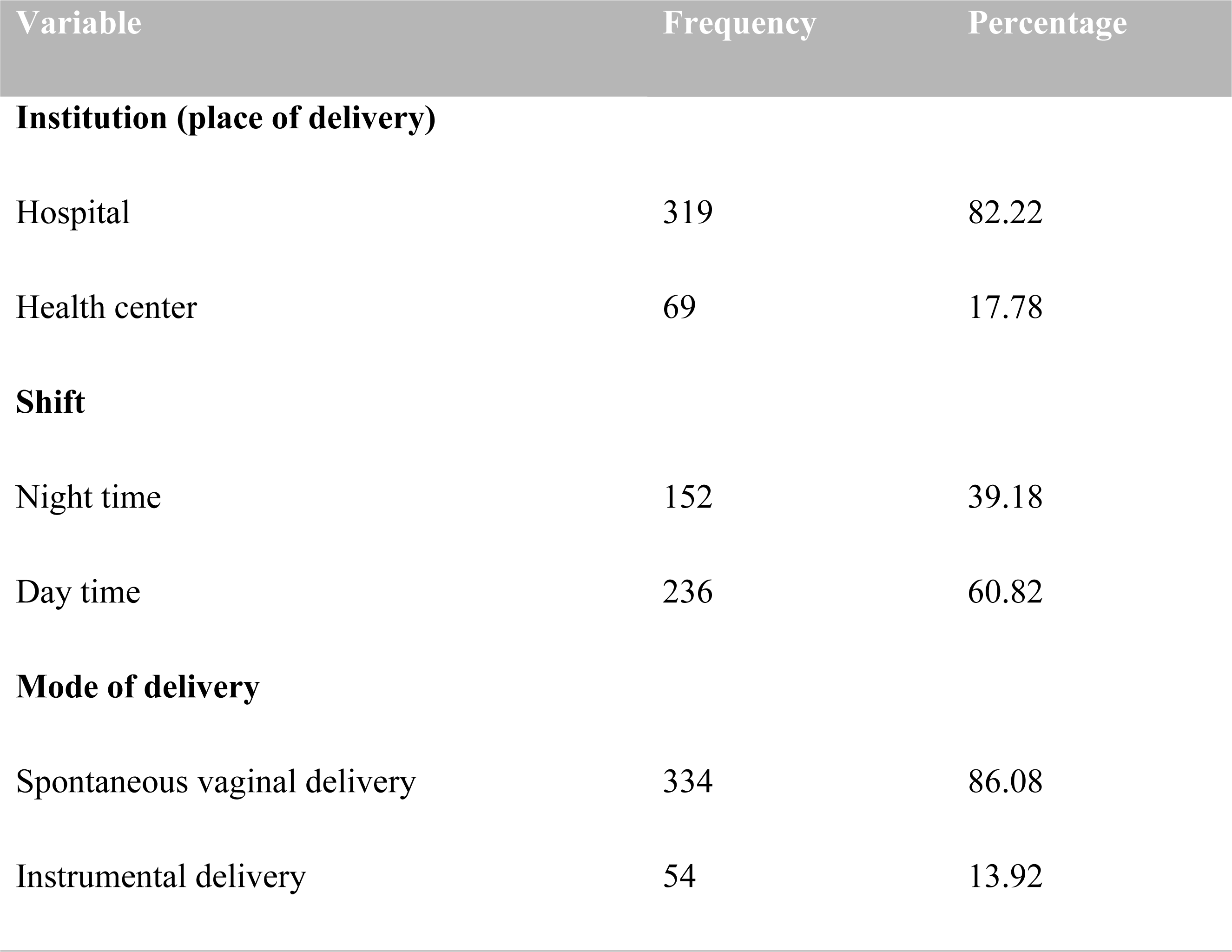
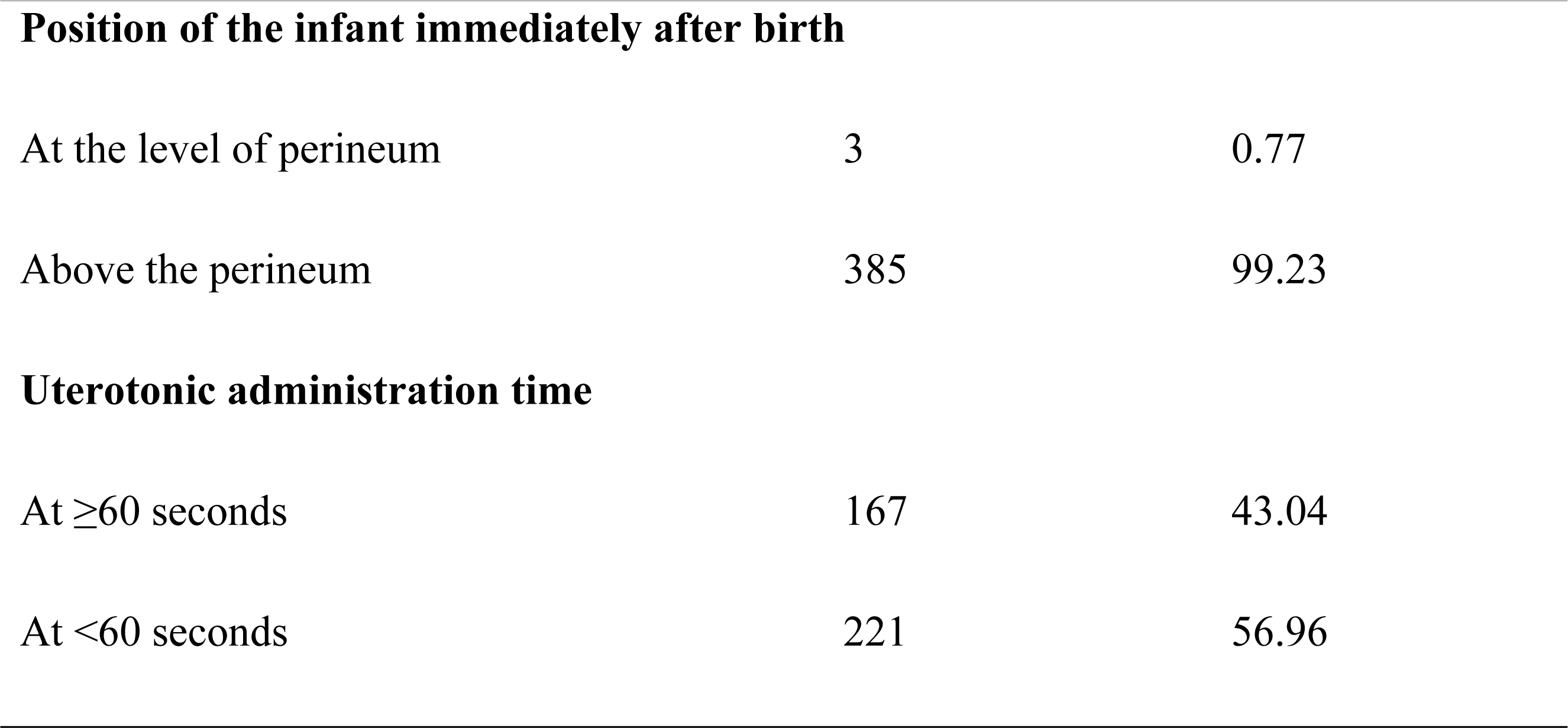
Labor delivery characteristics among women who gave birth in Debremarkos town public health institutions, north Ethiopia, 2022/23, (n=388)

### Obstetrics complications

Regarding obstetric complications, there were 53 (13.66%) induction or augmentation, 15.8 (3.87%) antepartum hemorrhage, 111 (28.61%) MSAF during labor delivery, 4 (1.03%) chorioaminoitis, and 13 (3.35%) preeclampsia or eclampsia. Women who gave birth with an episiotomy and a perineal tear occurring were 10 (2.5%), and 73 (18.81%), respectively (**Fig.3**).

**Fig 3.**
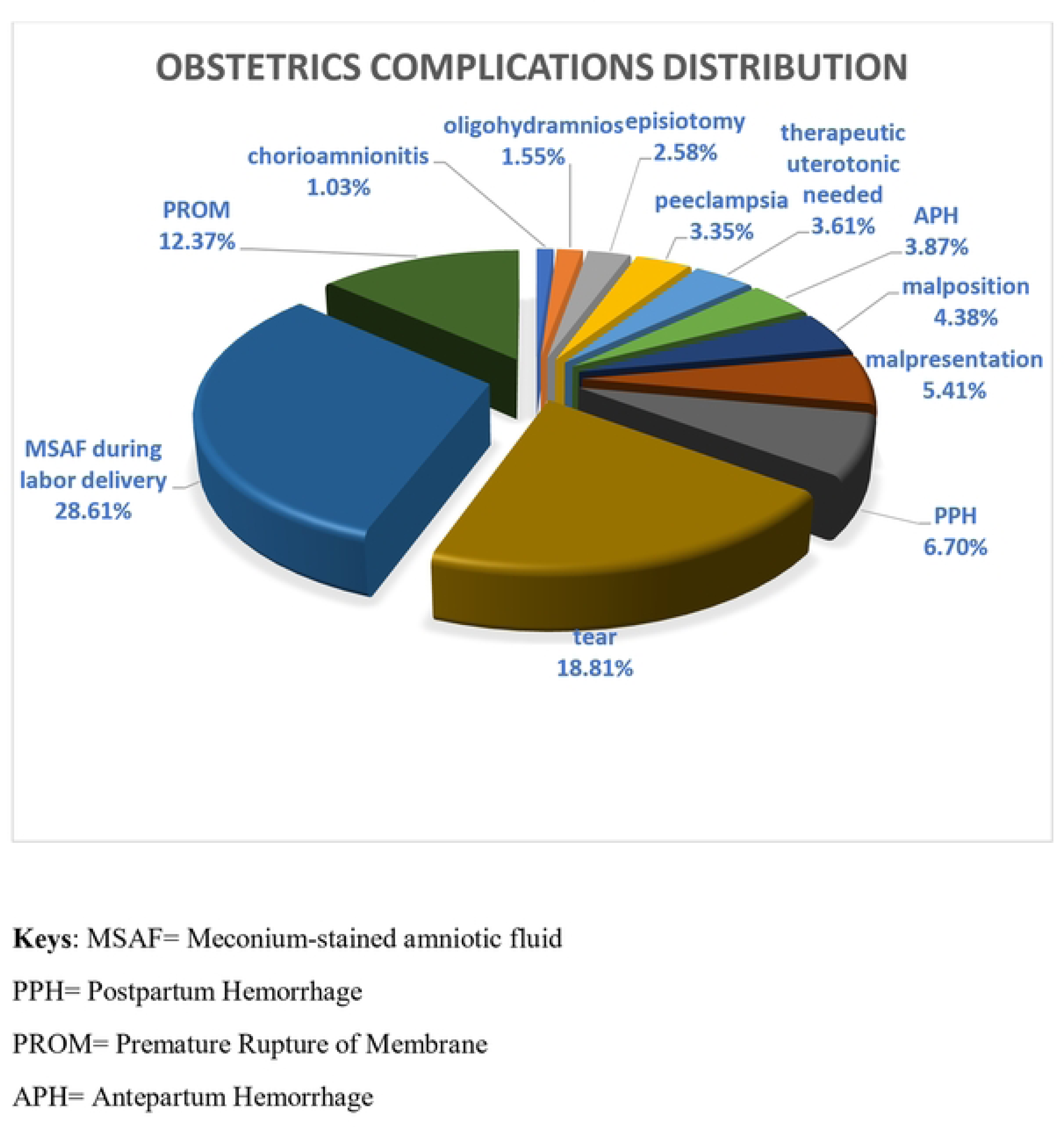
Distribution of Obstetrics complication among women who gave birth in Debremarkos to town public health institutions, north Ethiopia, 2022/23, (11=388)

### Professional factors

Midwives were cord clampers in 324 (83.51%) births (**Fig. 4**). There was teamwork in 323 (83.25%) deliveries, and the maximum number of HCPs in the team was 4. And there was an HCP dedicated to giving ENC in 328 (84.54%) cases.

**Fig 4.**
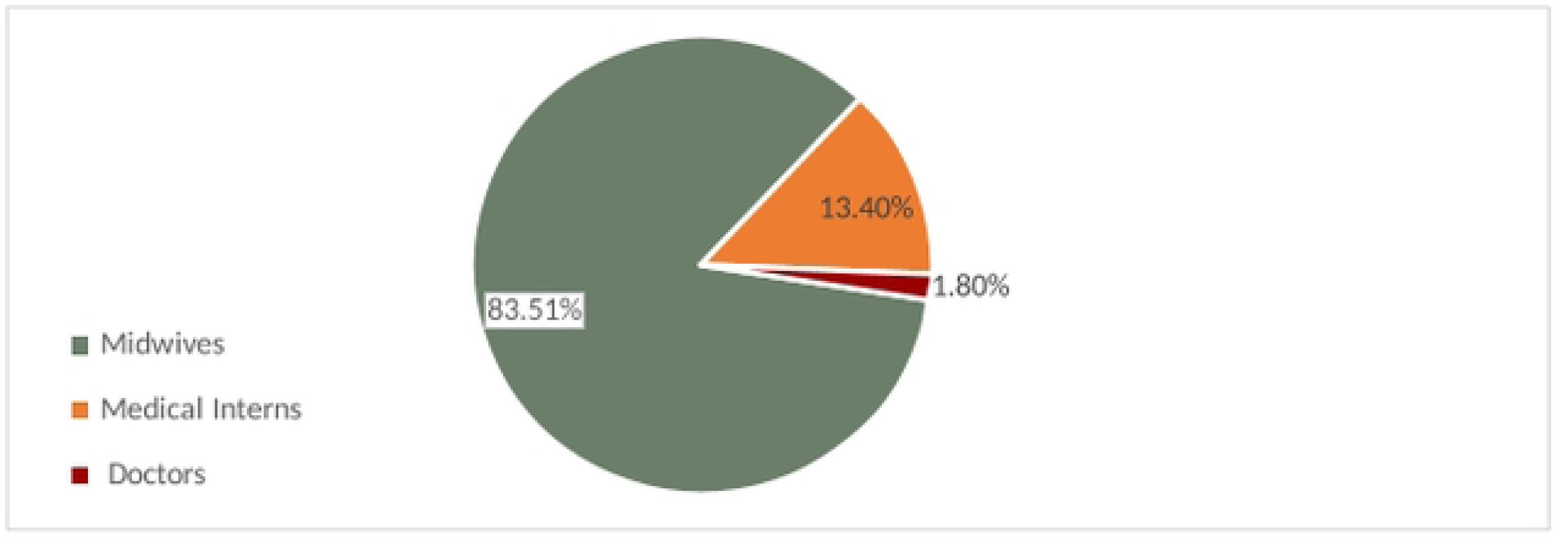
Professional difference of cord clampers in Debremarkos town public health institutions, north Ethiopia, 2022/23, (11=388)

### Umbilical cord clamping practice

The proportion of mothers who have received a delayed cord clamp (≥60 seconds) for their babies was 53.09% (95% CI: 48.09–58.03%). The range of umbilical cord clamping times in this study’s data set was 8 seconds to 187 seconds. The mean cord clamp time was 67.85 (SD 39.9) seconds (**Fig. 5**). Three of the 11 first-born twins and 5 of the 14 second-born twins had their umbilical cords clamped at or after 60 seconds of birth. Delayed cord clamp was received in 198 (53.66%) cephalic presentations and 4 (28%) breech presentations.

**Fig 5.**
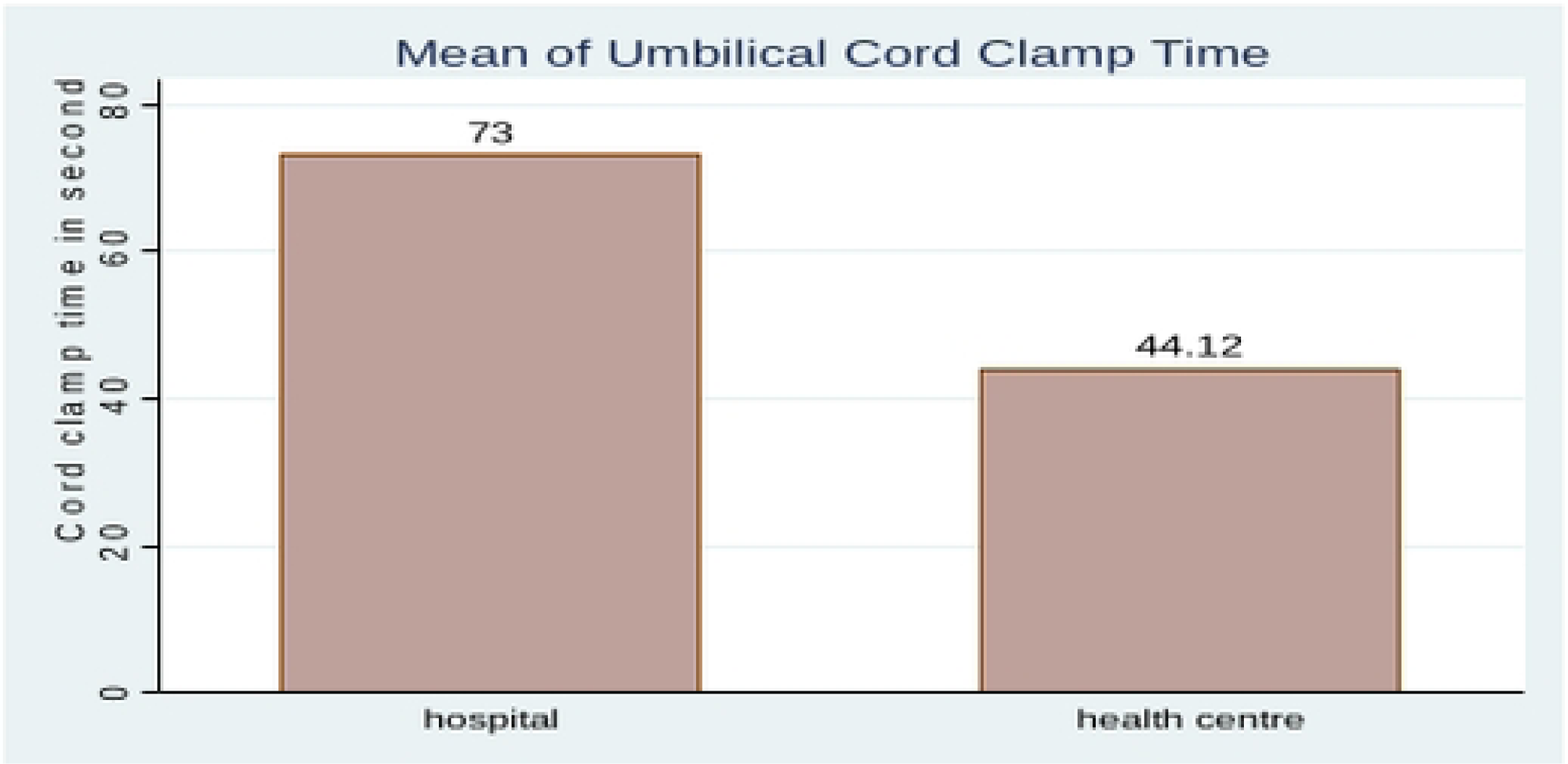
The n1ean cord clan1p time in Debrernarkos town public health institutions, north Ethiopia, 2022/23.

### Factors associated with the timing of umbilical cord clamp

#### Bivariable analysis

Findings from bivariable logistic regression showed that parity, maternal Rh status, gestational age, the need for resuscitation, place of delivery, mode of delivery, uterotonic for AMTSL administration time, cord milking, induction/augmentation, PROM, MSAF during labor delivery, perineal tear, the professional difference of the cord clamper, teamwork, and the presence of personnel dedicated to giving ENC were found to be associated with delayed cord clamp with a p-value of <0.2.

#### Multivariable analysis

In multivariable logistic regression parity, maternal Rh status, place of delivery, uterotonic administration time, and a professional difference in cord clamper were significantly associated with delayed cord clamp with a p-value of 0.05. women who gave birth at the hospital were 2.47 times more likely to receive DCC as compared to those who gave birth in the health centers (AOR = 2.47, 95% CI: 1.21-5.03). likewise, women who were attended by medical interns were 2.47 times more likely to receive DCC as compared to those who were attended by midwives (AOR = 2.47, 95% CI: 1.29-5.41). Similarly, among women to whom uterotonic for AMTSL was administered within greater than 60 seconds of giving birth, the odds of receiving DCC were 10.36 times greater than their counterparts (AOR = 10.36, 95% CI: 6.02-17.84). Rh-negative mothers were 3.91 times more likely to receive DCC than those who were Rh-positive (AOR = 3.91, 95% CI: 1.40-10.95). The odds of receiving DCC for their baby were 46% less likely among multipara women as compared to primipara women (AOR = 0.54, 95% CI: 0.32-0.93) (**Table 4**).

**Table 4.**
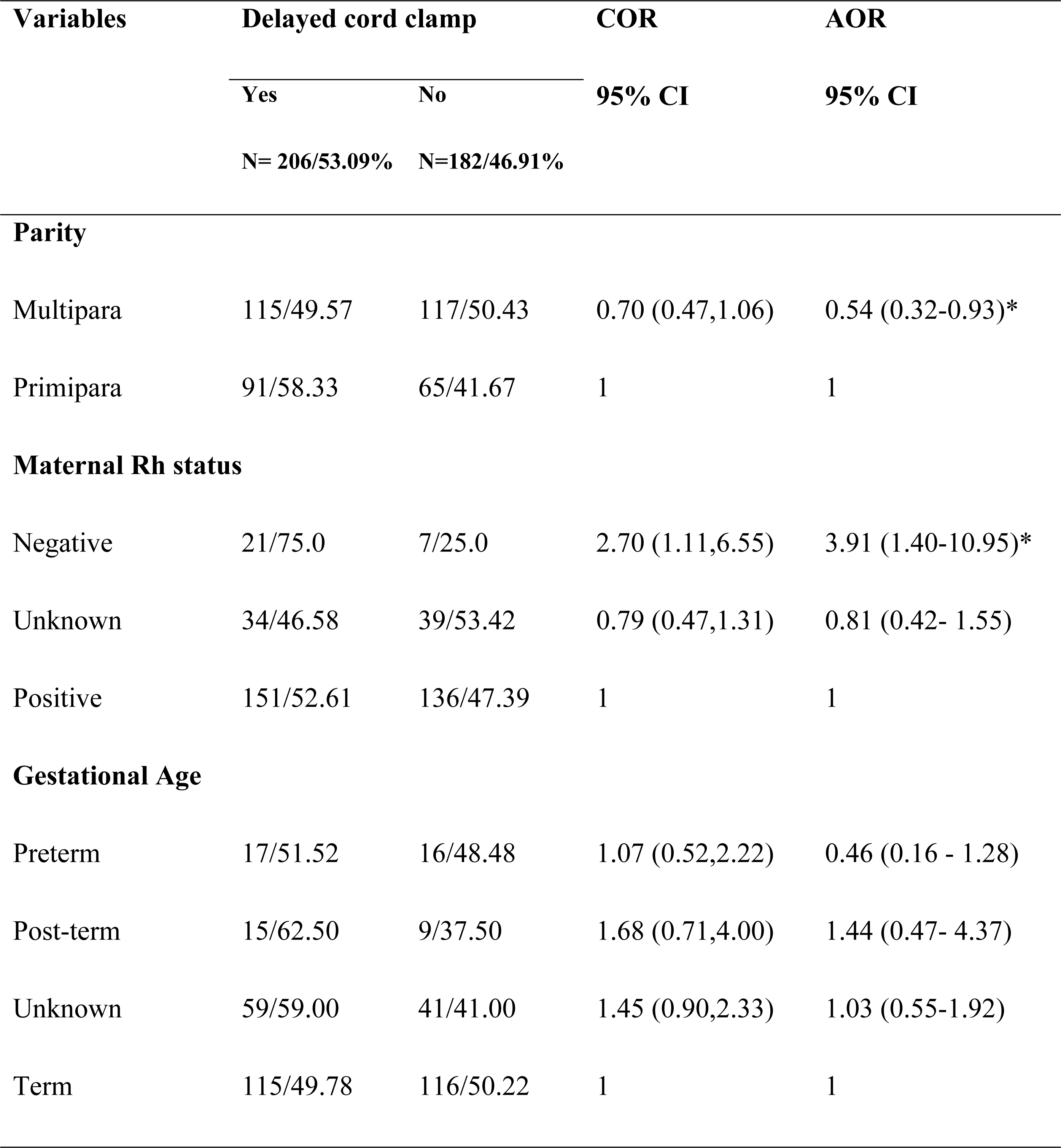
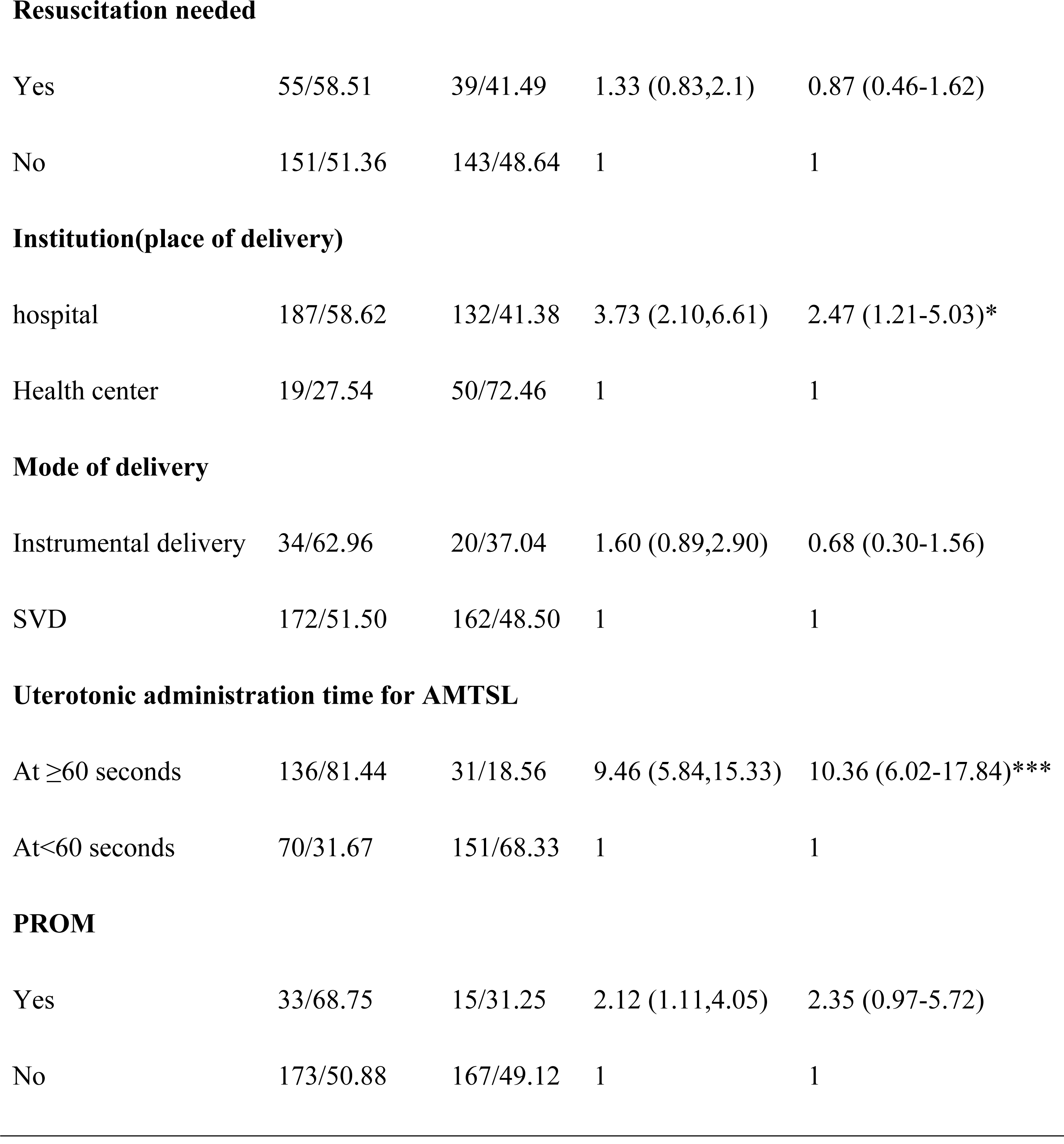
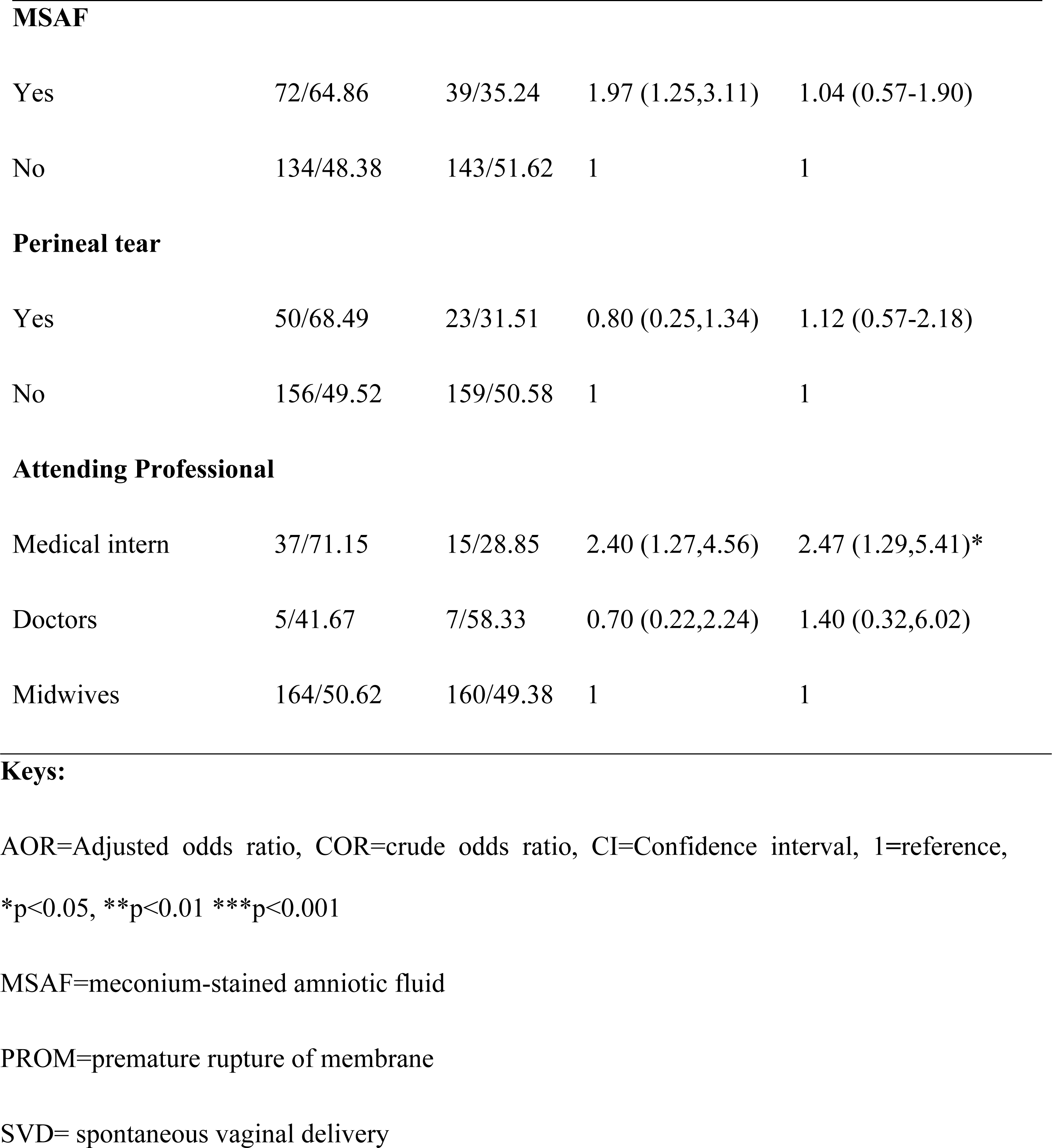
Bivariable and Multivariable analysis to identify factors associated with timing of umbilical cord clamping among women who gave birth in Debremarkos town public health institutions, North Ethiopia, 2022/23(n=388)

## Discussion

In this study, the percentage of delayed cord clamps (DCC) was found to be 53.09% (95% CI: 48.09-58.03%). This finding is comparable to the findings of studies conducted in Nepal (48%) (20) and Brazil (53.7%) (29). It is also in accordance with a study conducted in five public hospitals in Bangladesh, Nepal, and Tanzania, which shows that 47.3-73% of the newborns received DCC (32).

On the other hand, this finding is higher as compared to the study conducted in California (23%) (18). The disparity might be because the newborns in this study included all gestational ages greater than 28 weeks and also healthy newborns, whereas the study done in California included preterm and acutely ill infants admitted to neonatal intensive care units (18).

Regarding factors associated with delayed cord clamping, the place of delivery was found to be an important factor associated with DCC. In this study, women who gave birth at the hospital were 2.47 times more likely to receive DCC as compared to those who gave birth in the health centers (AOR = 2.47, 95% CI: 1.21-5.03). The ability to provide delayed umbilical cord clamping may vary among institutions and settings; decisions in those circumstances are best made by the team caring for the mother-infant pair (4). This can be explained by the fact that there are more care providers with higher education in hospitals than in health centers. As it is a teaching hospital, the hospital staff has more opportunities to be exposed to updates. Teaching hospitals are often synonymous with academic medical centers, which are expected to provide evidence-based care.

Regarding professional factors, professional differences were found to be significantly associated with DCC. In this study, the odds of DCC were 2.47 times higher among women who were attended by medical interns as compared to those who were attended by midwives (AOR = 2.47, 95% CI: 1.29-5.41). This can be explained by the fact that medical interns are expected to provide evidence-based care because they are actual students and have more access to updated and evidence-based information.

Prophylactic uterotonic administration time during the third stage of labor management was found to be another important factor significantly associated with DCC. Among women to whom uterotonic was administered at or after 60 seconds of giving birth, the odds of receiving DCC were 10.36 times greater than their counterparts (AOR = 10.36, 95% CI: 6.02-17.84). The reason may be related to the evidence that uterotonic drugs alter placental transfusion speed and duration (33). Thus, a delay in administering uterotonic during delayed cord clamping in this study might be necessary to avoid this uterotonic effect.

Maternal Rh status was found to be another factor associated with DCC. The likelihood that Rh-negative mothers would receive DCC was 3.91 times greater than that of Rh-positive mothers (AOR = 3.91, 95% CI: 1.40-10.95). This result does not fit the theory that DCC is not the usual norm due to fear of the potential risk of sensitization, hyper-bilirubinemia and exacerbation of anemia in these cases. The finding could be explained by a bias in estimation related to overestimations of short duration and underestimations of longer duration (34,35).

Furthermore, the odds of receiving DCC for their baby were 47% less likely among multiparous women as compared to primiparous mothers (AOR = 0.54, 95% CI: 0.32-0.93). This could be explained by the fact that multiparity increases the incidence of medical and obstetric complications, so earlier clamping of the umbilical cord may be required to manage complications, i.e., to provide essential care like newborn resuscitation. Supporting this, in this study, DCC was less likely in newborns who needed resuscitation. However, this result was not statistically significant. This is also supported by another result of this study. i.e., resuscitation was needed in 38.69% of babies born to mothers with obstetric complications, whereas it was only 8.99% among babies born to mothers without obstetric complications.

There are some limitations to this study that need to be addressed. Firstly, the healthcare providers did not know specifically that they were observed for their cord clamping practice, even though they were aware that they were observed. Therefore, the Hawthorne effect was reduced using participatory observation. Secondly, the data in this study cannot identify the reasons why delayed cord clamping was undertaken or not. And these data lack specific healthcare provider-related factors.

## Conclusion

Half of the newborns got delayed umbilical cord clamping, however the result is still unsatisfactory since the recommendations for DCC extended to all newborns, including those who do not require intensive care. According to this observational study, delivery attended by a medical intern, administering uterotonic for AMTSL at greater than 60 seconds of delivery, maternal Rh-negative status, and delivery at the hospital were found to contribute to delayed cord clamp. Whereas, multiparity was negatively associated with delayed cord clamping.

Health and educational bureaus should better design programs and strategies to increase delayed cord clamping practice. Healthcare providers should better consider the quantifiable benefits of delayed cord clamping and advocate for major adherence to DCC guidelines. Moreover, qualitative studies are needed to recognize potential barriers to the use of DCC and identify factors that would help facilitate a change in clinical practice and to explore the experience and perception of healthcare providers about the timing of cord clamping.

## Data Availability

Datasets are available from the corresponding author upon reasonable request.

## Abbreviations

AMTSL: Active Management of Third Stage of Labor
AOR: Adjusted Odds Ratio
CI: Confidence Interval
COR: Crude Odds Ratio
DCC: Delayed Cord Clamping
ECC: Early Cord Clamping
ENC: Essential Newborn Care
IDA: Iron Deficiency Anemia
NICU: Neonatal Intensive Care Unit
WHO: World Health Organization

## Consent for publication

Not applicable.

## Availability of data and materials’

Datasets are available from the corresponding author upon reasonable request.

## Authors’ contribution

BBW, BAK and EAC developed the concept of the research, acquired the research proposal, and facilitated data collection. BBW carried out statistical analysis. BBW, BAK and EAC draft the manuscript. All authors critically reviewed, revised and approved the manuscript.

## Acknowledgment

We are grateful to the Debremarkos Specialized and Comprehensive Hospital, the four health centers, and the heads of the respective MCH units and staff for their cooperation. Our deepest gratitude also goes to data collectors, supervisors, and study participants for their involvement in the data collection process.

## References

1. Pasricha SR, Tye-Din J, Muckenthaler MU, Swinkels DW. Iron deficiency. Lancet [Internet]. 2021;397(10270):233–48. Available from: 10.1016/S0140-6736(20)32594-0

2. Bayer K. Delayed umbilical cord clamping in the 21st century: Indications for practice. Adv Neonatal Care. 2016;16(1):68–73.

3. Molla A, Egata G, Mesfin F, Arega M, Getacher L. Prevalence of Anemia and Associated Factors among Infants and Young Children Aged 6-23 Months in Debre Berhan Town, North Shewa, Ethiopia. J Nutr Metab. 2020;2020.

4. ACOG. Delayed umbilical cord clamping after birth. Committee Opinion No. 684. American College of Obstetricians and Gynecologists. Obs Gynecol. 2017;129(684):e5–10.

5. Lozoff B, Beard J, Connor J, Felt B, Georgieff M, Schallert T. Long-lasting neural and behavioral effects of iron deficiency in infancy. Nutr Rev. 2006;64(5 SUPPL. 1).

6. Ibrahim NO, Sukkarieh HH, Bustami RT, Alshammari EA, Alasmari LY, Al-Kadria HM. Current umbilical cord clamping practices and attitudes of obstetricians and midwives toward delayed cord clamping in Saudi Arabia. Ann Saudi Med. 2017;37(3):216–24.

7. World Health Organization (WHO). Guideline: Delayed umbilical cord and nutrition outcomes maternal and infant health clamping for improved WHO. World Heal Organ. 2014;

8. Leslie MS. Perspectives on Implementing Delayed Cord Clamping. Nurs Womens Health. 2015;19(2):164–76.

9. Qian Y, Ying X, Wang P, Lu Z, Hua Y. Early versus delayed umbilical cord clamping on maternal and neonatal outcomes. Arch Gynecol Obstet [Internet]. 2019;300(3):531–43. Available from: 10.1007/s00404-019-05215-8

10. Raju TNK. Timing of umbilical cord clamping after birth for optimizing placental transfusion. Curr Opin Pediatr. 2013;25(2):180–7.

11. Rabe H, Gyte GML, Díaz-Rossello JL, Duley L. Effect of timing of umbilical cord clamping and other strategies to influence placental transfusion at preterm birth on maternal and infant outcomes. Cochrane Database Syst Rev. 2019;2019(9).

12. Bryce E, Mullany LC, Khatry SK, Tielsch JM, Leclerq SC, Katz J. Coverage of the WHO’s four essential elements of newborn care and their association with neonatal survival in southern Nepal. BMC Pregnancy Childbirth. 2020;6:1–9.

13. Village EG. Neonatal Outcome Following Cord Clamping After Onset of Spontaneous Respiration. Pediatrics. 2014;134:265–73.

14. NICE. Guideline:Intrapartum care for healthy women and babies. Nice. 2021;(December 2014).

15. Aydogan Kirmizi D, Başer E, Demir Çaltekin M, Onat T, Kara M, Yalvac ES. Behaviors and Attitudes of Obstetricians in Turkey Related to Cord Clamping, Cord Milking, and Skin-To-Skin Contact. Cureus. 2021;13(7):1–8.

16. Singh N, Brammer D. Delayed cord clamping in infants born less than 35 weeks: A retrospective study. J Neonatal Perinatal Med. 2020;1–5.

17. Boere I, Smit M, Roest AAW, Lopriore E, Van Lith JMM, Te Pas AB. Current practice of cord clamping in the Netherlands: A questionnaire study. Neonatology. 2015;107(1):50–5.

18. Tran CL, Parucha JM, Jegatheesan P, Lee HC. Delayed Cord Clamping and Umbilical Cord Milking among Infants in California Neonatal Intensive Care Units. Am J Perinatol. 2020;37(2):151–7.

19. Devin J, Larkin P. Delayed cord clamping in term neonates: Attitudes and practices of midwives in Irish hospitals. Int J Childbirth. 2018;8(1):4–17.

20. Nelin V, Kc A, Andersson O, Rana N, Målqvist M. Factors associated with timing of umbilical cord clamping in tertiary hospital of Nepal. BMC Res Notes [Internet]. 2018;11(1):1–6. Available from: 10.1186/s13104-018-3198-8

21. Leslie MS, Erickson-Owens D, Park J. Umbilical Cord Practices of Members of the American College of Nurse-Midwives. J Midwifery Women’s Heal. 2020;65(4):520–8.

22. Ortiz-Esquinas I, Gómez-Salgado J, Pascual-Pedreño AI, Rodríguez-Almagro J, Ballesta-Castillejos A, Hernández-Martínez A. Variability and associated factors in the management of cord clamping and the milking practice among Spanish obstetric professionals. Sci Rep. 2020;10(1):1738.

23. Grobman WA, Yee LM. Evaluation of Introduction of a Delayed Cord Clamping Protocol for Premature Infants in a High-Volume Maternity Center. HHS Public Access. 2018;129(5):835–43.

24. Braddick L, Tuckey V, Abbas Z, Lissauer D, Ismail K, Manaseki-holland S, et al. International Journal of Gynecology and Obstetrics CLINICAL ARTICLE A mixed-methods study of barriers and facilitators to the implementation of postpartum hemorrhage guidelines in Uganda. Int J Gynecol Obstet. 2016;132:89–93.

25. Sacks E, Mehrtash H, Bohren M, Balde MD, Vogel JP, Adu-bonsaffoh K, et al. Articles The first 2 h after birth : prevalence and factors associated with neonatal care practices from a multicountry, facility-based, observational study. Lancet Glob Heal. 2021;9:72–80.

26. Pichler G, Cheung PY, Binder C, O’Reilly M, Schwaberger B, Aziz K, et al. Time course study of blood pressure in term and preterm infants immediately after birth. PLoS One. 2014;9(12):114504.

27. Grobman WA, Yee LM. Evaluation of Introduction of a Delayed Cord Clamping Protocol for Premature Infants in a High-Volume Maternity Center. PMC. 2018;129(5):835– 43.

28. Hewitt T, Baddock S, Patterson J. Timing of cord clamping : An observational study of cord clamping practice in a maternity hospital in Aotearoa New Zealand. New Zeal Coll Midwives Journa. 2022;(58):19–26.

29. Strada JKR, Vieira LB, Gouveia HG, Betti T, Wegner W, Pedron CD. Factors associated with umbilical cord clamping in term newborns*. Rev Esc Enferm USP. 2022;56:1–8.

30. (WHO) C for IO of MS (CIOMS) in collaboration with the WHO. International Ethical Guidelines for Health-related Research Involving Humans [Internet]. Biomedical Research. 2016. 1921–1931 p. Available from: http://www.sciencedirect.com/science/article/B6VC6-45F5X02-9C/2/e44bc37a6e392634b1cf436105978f01

31. Technology FMoSa. National Research Ethics Review Guideline. 2015. p. 56.

32. Tahsina T, Hossain AT, Ruysen H, Rahman AE, Day LT, Peven K, et al. Immediate newborn care and validation study. BMC Pregnancy Childbirth [Internet]. 2020;21(Suppl 1):1–17. Available from: 10.1186/s12884-020-03421-w

33. Impact S, No P. Scientific Impact Paper No. 14: Clamping of the Umbilical Cord and Placental Transfusion. Obstet Gynaecol. 2015;17(3):216–216.

34. Roy MM, Christenfeld NJS, Jones M. Actors, observers, and the estimation of task duration. Q J Exp Psychol. 2013;66(1):121–37.

35. Lejeune H, Wearden JH. Vierordt’s the Experimental Study of the Time Sense (1868) and its legacy. Eur J Cogn Psychol. 2009;21(6):941–60.

